# Contrast-enhanced ultrasound for assessing tissue perfusion and predicting functional recovery following acute traumatic spinal injury: translation from rat model to humans

**DOI:** 10.1101/2024.01.04.24300837

**Authors:** Zin Z. Khaing, Jannik Leyendecker, Jennifer N. Harmon, Sananthan Sivakanthan, Lindsay N. Cates, Jeffrey E. Hyde, Melissa Krueger, Robb W. Glenny, Matthew Bruce, Christoph P. Hofstetter

## Abstract

Traumatic spinal cord injury (tSCI) leads to an immediate loss of neurological function, with its recovery being difficult to predict in the acute phase. Here, we developed a contrast-enhanced ultrasound (CEUS) imaging biomarker to quantify the intraspinal vascular disruption after tSCI. In rodent thoracic tSCI, CEUS revealed a perfusion area deficit (PAD) which increased with injury severity (p = 0.001). The PAD size significantly correlated with hindlimb locomotor function at 8 weeks post injury (R^2^ = 0.82, p < 0.001). Additionally, we calculated a spinal perfusion index (SPI) comparing the amount of perfused tissue at the injury center to that in injury periphery. Our experiments demonstrated that SPI decreased in more severe injuries and correlated significantly with hindlimb locomotor function at 8 weeks post injury (R^2^ = 0.83, p < 0.001). Subsequently, we demonstrated the feasibility of intraoperative CEUS imaging in 20 patients with acute tSCI. A hyper-perfusion pattern was commonly seen in cervical motor-incomplete tSCI, while necrosis penumbra pattern was associated with motor-complete cervical or thoracic tSCI. We measured both PAD and SPI and detected statistically significant differences between motor-complete and motor-incomplete patients. In our patient cohort, SPI exhibited a strong predictive capacity for functional recovery at 6 months (R^2^ = 0.79, p < .001) compared to PAD. In conclusion, our study suggests that an intraoperative CEUS-derived biomarker holds promise for predicting injury severity and chronic functional outcome after tSCI. Larger clinical studies are needed to better assess the reliability of the proposed CEUS-derived biomarker and its prognostic capacity.

**One Sentence Summary:** This paper introduces a novel biomarker utilizing contrast-enhanced ultrasound in humans and rats that accurately predicts injury severity after traumatic spinal cord injury based on tissue perfusion.

## Introduction

Traumatic spinal cord injury (tSCI) results in an immediate loss of neurological function, with concomitant functional, psychological, and financial consequences for both the affected individuals and their families ^1, 2^. In the United States, the annual incidence of tSCI is 26 new cases per 100,000 individuals, which is double the global incidence rate ^3^. Currently, more than a quarter million Americans are living with tSCI ^4^, resulting in estimated annual treatment costs of $9.7 billion ^5^.

Aghast by the traumatic event, the tSCI victim and their families’ initial conversations with the medical team consistently revolve around the anticipated functional recovery. In clinical practice, injury severity and long-term functional outcome is prognosticated dependent on the neurological examination according to the International Standards for Neurological Classification of Spinal Cord Injury (ISNCSCI) at presentation to the trauma hospital ^6, 7^. ISNCSCI has been demonstrated to be a valid, reliable, and responsive tool to assess adults with tSCI ^8^. Several studies have confirmed *high intra-rater and inter-rater reliability* ^9, 10^. However, the impact of timing of the ISNCSCI examination onto its prognostic capacity remains controversial ^11^. Historically, the ISNCSCI was obtained when the patient was transferred from the acute hospitalization to an inpatient rehabilitation facility approximately 1 month after tSCI and served as a baseline for predicting long-term outcome ^12–15^. However, the patients’ demand for early prognostication and the relatively short therapeutic window of pre-clinical neuroprotective interventions have motivated the utilization of the ISNCSCI exam performed at arrival at the trauma center ^16–18^. The need for this early prognostic biomarker is substantiated by the finding that spinal cord decompression within 24 hours following tSCI improves sensorimotor recovery, whereas surgery 24–36 h after injury is associated with a steep decline in recovery ^19^.

Several factors might contribute to the limited prognostic capacity of the ISNCSCI examination in the acute phase after injury. First, even mild tSCI with supple impingement of the spinal cord can cause transient spinal cord ischemia with temporary loss of motor and sensory function also referred to as neurapraxia, especially in very early examinations ^20^. *Second,* injury expansion within the first 72 hours might be another contributor to *account for differences in the predictive capabilities of the* ISNCSCI examination within the first 24 hours compared to 72 hours post injury. Within the first hours after injury, the hypoperfused spinal cord injury center enlarges in rodents and humans following tSCI^21^. ^22^. Lastly, an ISNCSCI examination is impossible in patients with concomitant traumatic head injury, multi-system trauma, drug intoxication, or pharmacological sedation. In summary, several studies suggest that the prognostic capacity of the ISNCSCI examination is limited within the first 72 hours after tSCI ^23–26^.

Given the significant demand for early prognostication, both neurochemical and imaging markers have been developed to serve as tSCI biomarkers. Cerebral spinal fluid (CSF) collected 24 hours after tSCI contains inflammatory markers and structural proteins that accurately classify tSCI severity and predict neurological recovery ^27^. However, collection of CSF requires a time consuming lumbar puncture and laboratory analysis which is not standardized between centers. Analysis of MRI imaging parameters such as intramedullary lesion length, hemorrhage, and spinal cord swelling measured on acute posttraumatic magnetic resonance imaging (MRI) have shown promise as possible predictive biomarkers ^28–30^. However, analysis of spinal cord parameters on MRI can be challenging in particular with the spinal cord compressed and the spinal columns misaligned. Here we propose contrast enhanced ultrasound (CEUS) as a promising quantitative imaging biomarker for acute tSCI. This technique depicts blood flow in the micro- and macrocirculation ^21, 31–34^, disruption of which constitutes a hallmark pathophysiological event in tSCI evolution ^35^. Importantly, its high spatial resolution allows for quantification of the perfusion deficit area ^34^. Furthermore, CEUS imaging provides the surgeon with instant feedback regarding adequacy of spinal cord decompression ^36^.

Our current study solidifies the key role of vascular disruption in the pathophysiology of tSCI. We report the development of two complementary CEUS-based biomarkers: perfusion deficit area (PAD) which quantifies the amount of damaged hypoperfused spinal cord tissue and spinal perfusion index (SPI) which quantifies the amount of spared perfused spinal tissue at the injury center. Collection of the CEUS-based biomarker in our rodent tSCI revealed an injury severity-dependent homogenous pattern of vascular disruption. In rat tSCI, both PAD and SPI were excellent predictors of injury severity and functional outcome. In contrast, disruption of vascular perfusion was heterogenous in our patient cohort with both hyper-perfusion and necrosis penumbra pattern detected. Our preliminary data suggest that instant intraoperative SPI collection is feasible and might be a valuable novel type of biomarker for tSCI severity in human tSCI.

## Results

### tSCI causes disruption of spinal cord blood flow

Disruption of spinal cord parenchymal perfusion, the basis for our proposed quantitative imaging biomarker, was first studied utilizing three independent techniques – ultrafast ultrasound imaging, microsphere deposition assay, and histological vessel patency analysis. Adult rodents were subjected to a moderate thoracic tSCI (150 kDyn). After tSCI, CEUS midline sagittal images were acquired. CEUS Doppler revealed disruption and displacement of intrinsic spinal cord blood vessels at the injury center (Fig 1A). Examination of blood flow changes within the microcirculation revealed a perfusion area deficit centered in the injury center measuring on average 1.32 ± 0.43 mm^2^. We then analyzed the relative blood volume in rostrocaudal spinal cord segments centered over the injury center. A 2/3 reduction of blood volume was seen at the injury center extending 3 mm rostrocaudal. Next, microsphere deposition assay was carried out to corroborate disruption of tissue perfusion in the acutely contused spinal cord (Fig 1B). Disruption of tissue perfusion was complete in the gray matter. Similar to the findings with CEUS, a substantial decrease of white matter blood flow was noted in an approximately 3 mm rostrocaudal spinal cord segment. Analysis of lectin-labeled histology confirmed extensive disruption of intrinsic spinal cord blood vessels at the injury site (Fig. 1C). Thus, in the injury center, the density of intrinsic spinal cord blood vessels was diminished by 38.6 ± 9.63% in the grey matter and by 39.0 ± 12.0% in the white matter. Of the remaining blood vessels, 28.9 ± 13.9% remained patent in the gray matter and 54.6 ± 17.3% in the white matter. Loss of spine vessel patency was most pronounced in the central gray matter and dorsal white matter tracts.

**Fig 1.**
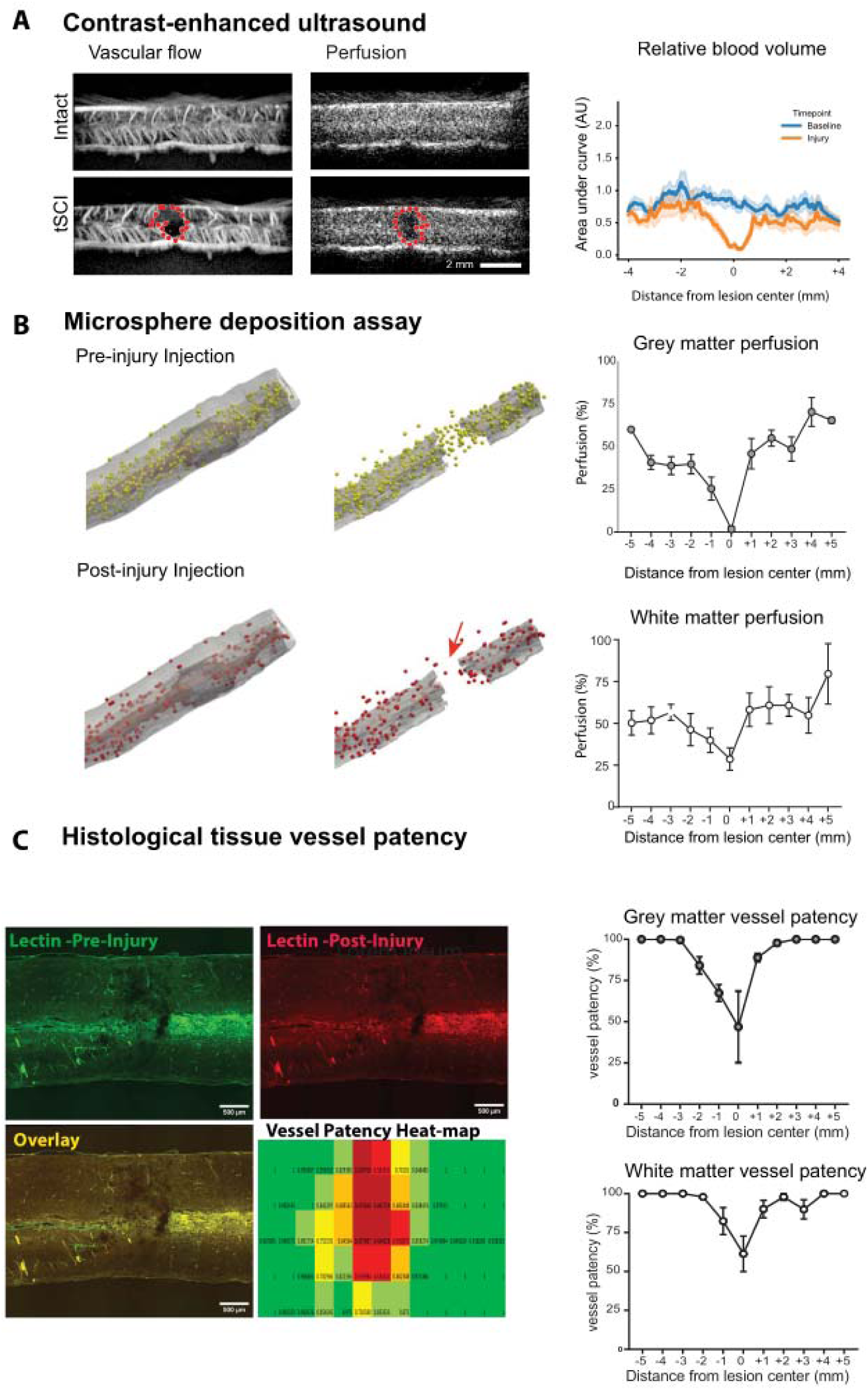
tSCI causes disruption of spinal cord blood flow and perfusion. (A) Contrast-enhanced ultrasound (CEUS) midline sagittal images of the thoracic rat spinal cord. Moderate severity tSCI results in disruption/displacement of intrinsic spinal cord blood vessels and loss of tissue perfusion at the injury center (red dotted line). Line graphs depict the effect of tSCI on blood volume (area under the curve) in rostro-caudal spinal cord segments centered on the injury center. (B) Representative microsphere deposition assays for both pre-injury (yellow beads) and post-injury (red) injections. Almost complete loss of tissue perfusion is detected in the grey matter at the injury center. Hypoperfusion of the white matter perfusion is noted spanning approximately 3 mm in rostral-caudal direction. (C) Pre- and post tSCI lectin vascular labeling demonstrates both disruption and occlusion of intrinsic spinal cord blood vessels at the injury site. The heat map demonstrates loss of vessel patency predominately in the central gray matter and dorsal white matter tracts at the injury site. (Heatmap vessel patency: dark red <50%, light red <60%, orange <70%, yellow<80% and light green <90% patency. The graphs demonstrate loss of vessel patency at the injury site more pronounced in the gray matter compared to the white matter. An approximately 3 mm segment of the spinal cord centered at the injury site depicts loss of vessel patency.

### PAD correlates with injury severity in rodent tSCI

We then studied how the injury severity influences the extent reduced blood flow in the spinal cord parenchyma (Fig. 2A). Rodents were subjected to mild, moderate, or severe tSCI resulting in graded degrees of hindlimb locomotor dysfunction (Fig. 2B – C). Some degree of spontaneous locomotor and sensory recovery, measured by open-field locomotor test and ladder walk performance was detected in animals of all three severity groups. Various injury severity groups exhibited significantly different behavioral dysfunction during the 8 week follow up duration. In all animals CEUS was performed acutely following tSCI. The perfusion area deficit (PAD) measured 0.41 ± 0.10 mm2, 1.32 ± 0.43 mm2, and 2.87 ± 0.27 mm2 in mild, moderate, and severe tSCI, respectively (p = 0.001, Fig. 2D). The extent of the PAD increased in both rostrocaudal and dorsoventral directions with greater injury severity (Fig. 2D). The PAD size measured on acute CEUS showed a very strong statistically significant correlation with hindlimb locomotor deficit at both 2- and 8-weeks post injury (R^2^ = 0.80, p < 0.001 and R^2^ = 0.82, p < 0.001 respectively; Fig. 2E). We also detected statistically significant correlations of the PAD with 2- and 8-weeks post injury ladder walk data (R^2^ = 0.60, p < 0.01 and 0.65, p < 0.001 respectively; Fig. 2F) as well as the extent of spared myelin at the epicenter at 8wpi (R^2^ = - 0.72, p < 0.001; see Supplementary data). Combined, these data strongly suggest that PAD measured acutely after SCI has a strong predictive capacity for chronic functional deficits in rodent tSCI.

**Fig 2.**
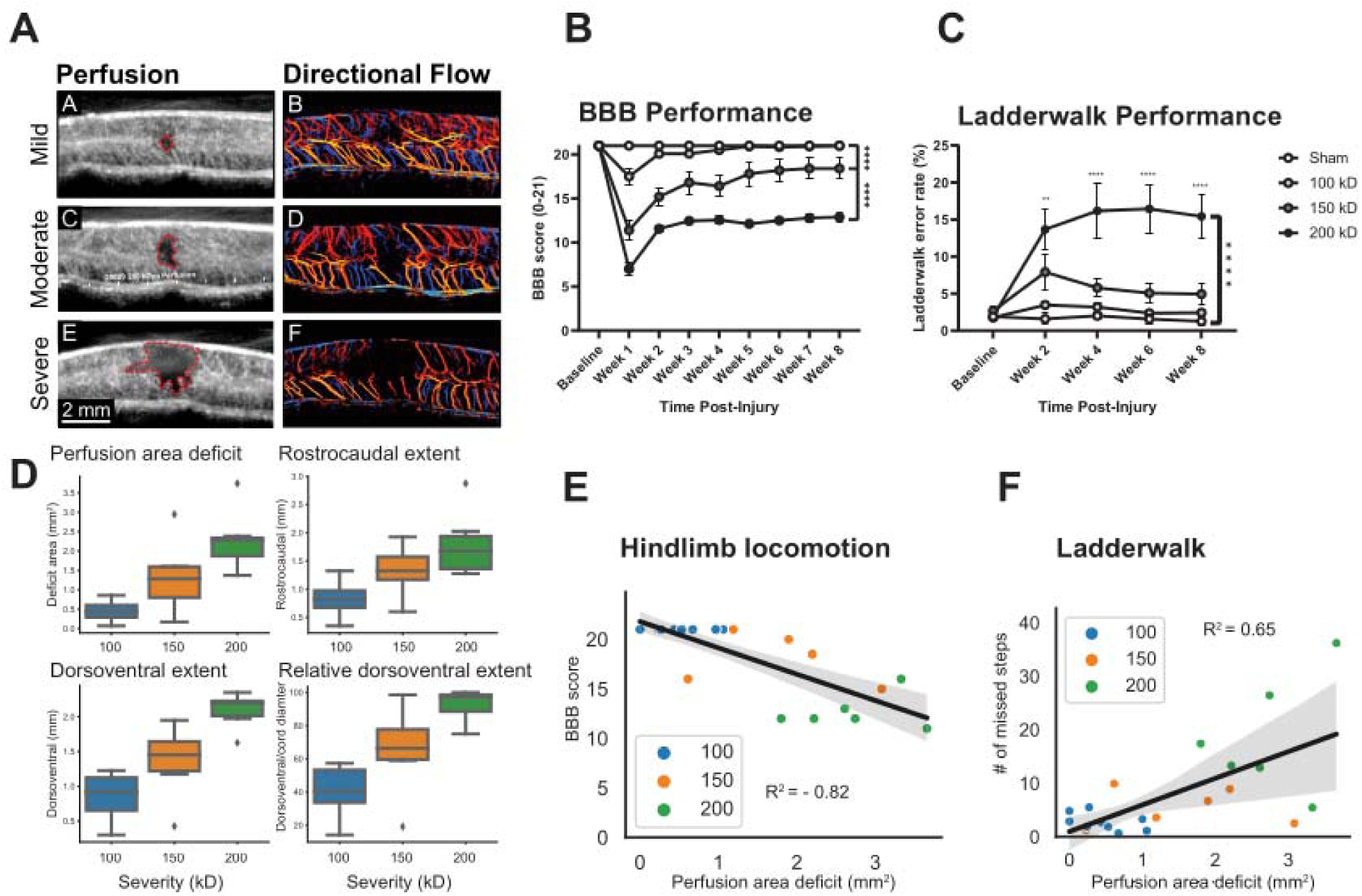
Perfusion area deficit (PAD) correlates with functional outcome in rodent tSCI. (A). Representative images of ultrafast CEUS Doppler depicting tissue perfusion (left panel) and super-resolution ultrasound localization imaging depicting directional flow (right panel) after a mild, moderate, or severe tSCI are shown. PAD is outlined by a red dotted line. Dorsally directed flow is color-coded in red, depicts ventrally directed flow (B - C) Behavioral assessments confirm increased levels of locomotor impairment with more severe levels of injury using both BBB and ladder walk testing. (D) With increasing injury severity, a significant increase in the PAD was measured. Similarly, the rostro-caudal, dorsoventral, and relative dorsoventral dimensions increase corresponding to the injury severity. (E - F). PAD measured acutely after injury correlates with chronic hindlimb locomotor recovery at 8 weeks post injury. Hindlimb locomotion is assessed by Basso, Beattie, and Bresnahan test (BBB) or ladder walk.

### Spinal perfusion index (SPI) correlates with functional outcome in rodent tSCI

As possible surrogate for spared tissue, we quantified residual perfused spinal cord parenchyma at the injury center. We designed a spinal perfusion index (SPI) as the contrast-inflow in the injury center normalized to the contrast-inflow in spared spinal cord parenchyma rostral and caudal to the injury (Fig. 3A) The SPI was calculated in all CEUS obtained acutely following tSCI. The SPI measured 0.89 ± 0.04, 0.51 ± 0.11, and 0.34 ± 0.06 in mild, moderate, and severe tSCI, respectively (Fig. 3B). The SPI in mild tSCI was statistically significantly higher compared to moderate (p = 0.01) and to severe (p < 0.001). The SPI determined on acute CEUS exhibited a very strong statistically significant correlation with hindlimb locomotor deficit at both 2- and 8-weeks post injury (R^2^ = 0.84, p < 0.001 and R^2^ = 0.83, p < 0.001 respectively; Fig. 3C). Similarly, we also detected statistically significant correlations of the SPI with 2- and 8-weeks post injury ladder walk data (R^2^ = 0.71, p < 0.001 and 0.58, p < 0.01 respectively; Fig. 3D) as well as the amount of spared myelin at the epicenter at 8wpi (R^2^ = - 0.68, p < 0.01; see Supplementary data). The SPI exhibited an extremely strong correlation with the PAD (R^2^ = - 0.90, p < 0.001; Fig. 3E) suggesting homogeneity of various severity rodent tSCI with regard to tissue damage and sparing.

**Fig 3.**
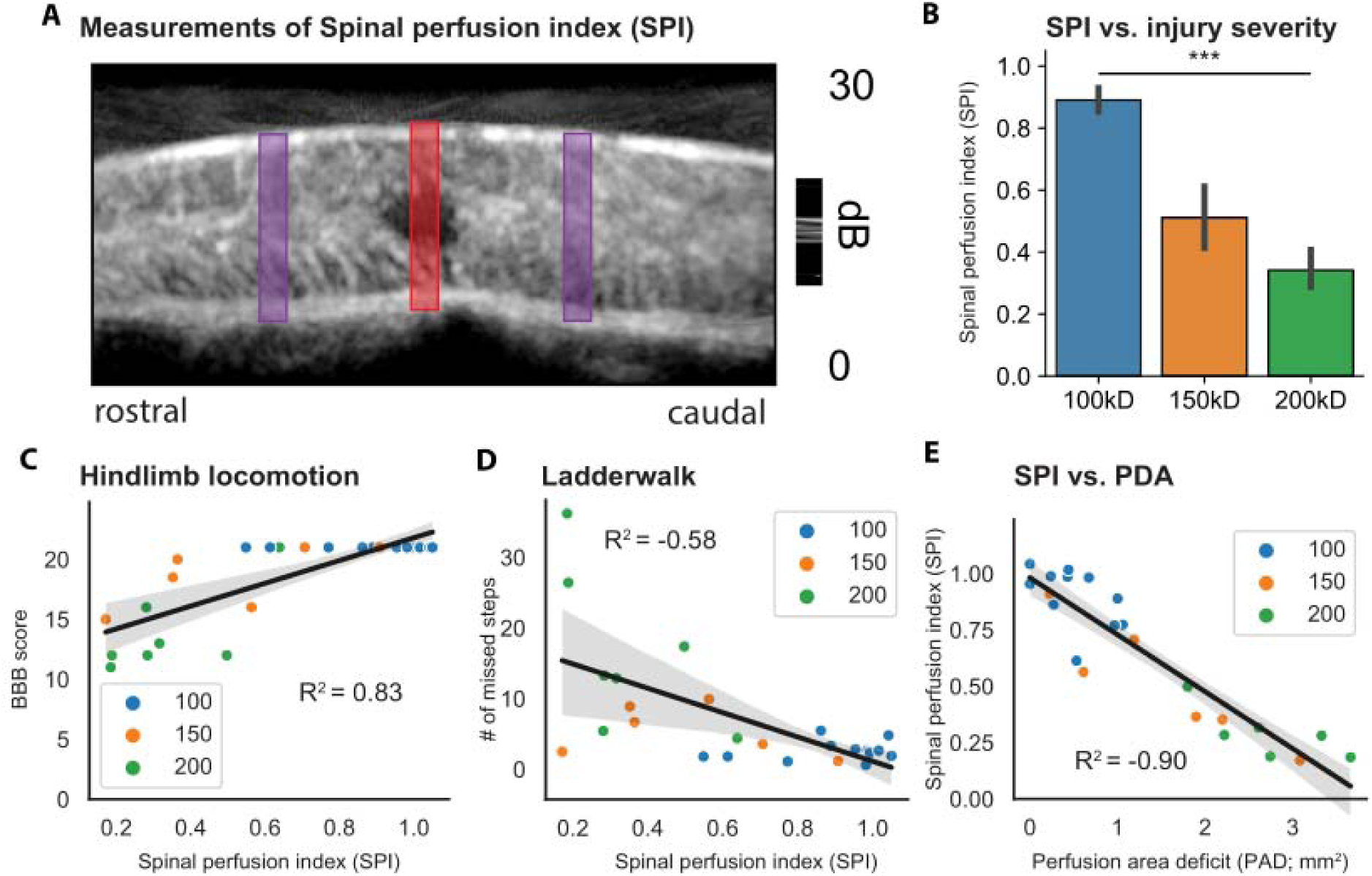
Spinal perfusion index (SPI) correlates with injury severity and functional outcome in rodent tSCI. (A). Example for Spinal perfusion index (SPI) measurement on CEUS study. The area under curve within the ROI at the injury center (red) is normalized to the average area under the curve in the rostral (green) and caudal (blue) ROI. (B) A minimal decrease of the SPI is detected in mild rodent tSCI, while a significant reduction is measured in animals with more severe injury. (C-D) SPI determined acutely after injury exhibits a strong correlation with the hindlimb locomotor score (BBB) as well as the number of missed steps on the ladder walk testing at 8 weeks post injury. (E) SPI and PAD show a very strong negative correlation in a rodent tSCI model.

### Feasibility to collect CEUS-based biomarker in acute tSCI patients

A total of 20 patients with acute tSCI who underwent posterior surgical spinal decompression and stabilization were included in our study (**Table 1**). The cohort was on average 58.9 ± 4.8 years old and consisted of 4 female and 16 male patients (Table 1). Motor vehicle accidents were the cause for tSCI in half of our patients followed by falls from ground level and falls from height. Most of the injuries were in the cervical spine (85.0%) while 3 patients suffered from injury to the thoracic spine. Complete neurological impairment (AIS grade A) was documented in 8 patients (40.0%) while the remaining patients suffered from incomplete tSCI. Patients underwent posterior surgical decompression and stabilization followed by intraoperative CEUS on average 17.6 ± 3.2 hours after the traumatic event (**Table 2**). The arthrodesis construct involved on average 5.3 ± 0.4 spinal segments and spanned the cervicothoracic junction in more than half of our patients. All patients underwent direct decompression of the contused spinal cord by removing on average 3.7 ± 0.4 laminae as well as stabilization of the involved spinal segments. The estimated blood loss was on average 440.0 ± 104.2 ml. Surgeries took on average 212.3 ± 18.2 minutes to complete. Prior to closing the surgical wound, all patients underwent CEUS imaging of the contused spinal cord. B-mode images revealed pathological hyperechogenicity at the site of tSCI in all patients. In mild tSCI, hyperechogenicity was typically confined to the central grey matter. An intravenous bolus of contrast agent was given, and contrast inflow was recorded on sagittal midline CEUS. Surgeries and CEUS were well tolerated. We did not encounter any intraoperative complications associated with surgeries or contrast-enhanced ultrasound imaging. Two patients required revision surgeries for surgical site infections.

**Table 1.** Patient demographics depicted as mean with standard error or absolute number with percentages for a total of 20 patients.

| **Parameter** | **Mean (SEM)** | **Frequency (%)** |
| --- | --- | --- |
| **Age (years)** | 58.9 ± 4.8 | - |
| **Sex** |  |  |
| Male | - | 16 (80%) |
| Female | - | 4 (20%) |
| **Mechanism** |  |  |
| MVC | - | 10 (50%) |
| Fall | - | 5 (25%) |
| Fall (height) | - | 5 (25%) |
| **Neurological level of injury** |  |  |
| Cervical | - | 17 (85%) |
| Thoracic | - | 3 (15%) |
| **ASIA admission** |  |  |
| A | - | 8 (40%) |
| B | - | 0 (0%) |
| C | - | 5 (25%) |
| D | - | 7 (35%) |
| **Time until MRI (hours; n=13)** | 7.77 (2-18) | - |
| **Rostrocaudal injury extension on MRI (cm)** | 2.92 (0.31) | - |

**Table 2.** Clinical details depicted as mean with standard error or absolute number with percentages for a total of 20 patients.

| **Parameter** | **Mean (SEM)** | **Frequency (%)** |
| --- | --- | --- |
| **Interval between trauma and surgery (hours)** | $17.56 \pm 3.2$ | - |
| <12 hours | - | 8 (40%) |
| < 24 hours | - | 15 (75%) |
| **Posterior segmental instrumentation (PSIF)** | $5.3 \pm 0.4$ levels | - |
| **Anatomical location PSIF** |  |  |
| Cervical | - | 6 (30%) |
| Cervicothoracic | - | 11 (55%) |
| Thoracic | - | 3 (15%) |
| **Laminectomy** | $3.7 \pm 0.4$ levels | - |
| **Surgery time** |  |  |
| Total | $212.3 \pm 18.2$ min | - |
| Per level | $41.91 \pm 3.57$ min | - |
| **Blood loss** |  |  |
| Total | $440.0 \pm 104.2$ ml | - |
| Per level | $88.23 \pm 21.34$ ml | - |
| **Duration of hospital stay** | - | 18 (2.92%) |
| **Discharge** |  |  |
| Home | - | 5 (25%) |
| Rehab | - | 12 (60%) |
| Death | - | 3 (15%) |
| Surgical site infections | - | 2 (10%) |

**Table 3.** Patient Outcomes for neurological status at various timepoints. Values are shown as absolute number with percentages for a total of 20 patients.

| **Parameter** | **Value (n)** |
| --- | --- |
| ASIA grade admission |  |
| A | 8 (40%) |
| B | 0 (0%) |
| C | 5 (25%) |
| D | 7 (35%) |
| ASIA grade mortality |  |
| A | 3 (60%) |
| B | 0 (0%) |
| C | 1 (20%) |
| D | 1 (20%) |
| ASIA grade discharge (n=17) |  |
| A | 7 (41.2%) |
| B | 0 (0%) |
| C | 0 (0%) |
| D | 10 (58.8%) |
| E | 1 (5.8%) |
| ASIA grade improvement | 12 (60%) |
| A | 0 (0%) |
| B | 0 (0%) |
| C | 3 (25%) |
| D | 9 (75%) |
| ASIA grade last Follow-Up (n=15) |  |
| A | 5 (33.3%) |
| B | 0 (0%) |
| C | 0 (0%) |
| D | 7 (13.7%) |
| E | 3 (20%) |

Analysis of contrast inflow on sagittal ultrasound studies revealed a perfusion deficit area (PAD) in 16 (80%) patients (Fig. 4A). All patients without discernable PAD were AIS D at admission. Patients with incomplete tSCI had a PAD that was significantly smaller compared to the PAD detected in complete tSCI (18.4 ± 9.3mm2 vs. 79.5 ± 13.1mm2; p < 0.01; Fig. 4B). PAD measured on acute intraoperative CEUS exhibits a correlation with the AIS score at last follow-up (R^2^ = 0.5, p < .001; Fig. 4C). ROC analysis revealed that a PAD cutoff of 37.4 mm^2^ predicted completeness of the spinal cord injury with an area under the curve of 0.917 (P < 0.001; Fig. 4D)

**Fig 4.**
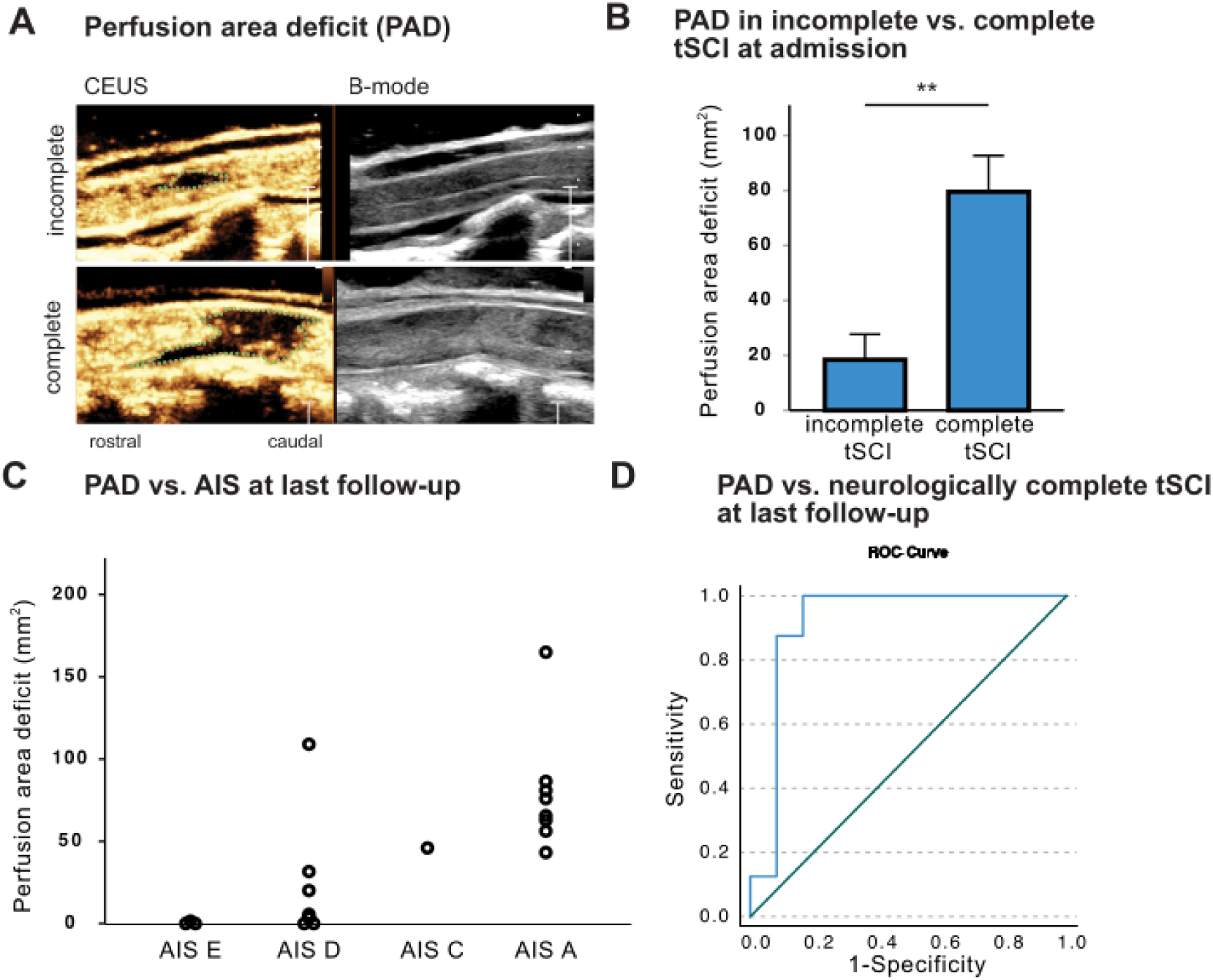
Assessment of the perfusion area deficit (PAD) in human tSCI. (A) Representative intraoperative ultrasound displaying tissue perfusion (left panel) and B-mode (right panel) images after surgical decompression of a motor incomplete (upper row) and motor complete (bottom row) human tSCI. The PAD is demarcated by a green dotted line. (B) Comparison of the PAD between admission exam motor incomplete (ASIA C-E) and motor complete (ASIA A-B) patients. A significantly greater perfusion deficit is detected in motor complete patients. (C) Scatterplot of tSCI patients depicting the PAD in relation to ASIA grade at last follow-up. Patients with better neurological outcomes have a lower perfusion area deficit. (D) Receiver-operating-characteristics of PAD vs. completeness of tSCI at last follow-up. A PAD cutoff of 37.4 resulted in an area under the curve of 0.917 (P < 0.001).

To quantify the amount of spared perfused spinal cord tissue at the injury center, we calculated the spinal cord perfusion index (SPI) in each patient (Fig. 5A). In contrast to rodent tSCI, patients with mild tSCI (AIS D) had pronounced hyperemia at the site of injury as evidenced by an average SPI of 1.37 ± 0.07. All hyperemic tSCI lesions were located in the cervical spinal cord. The intraoperative SPI exhibited a strong correlation with the AIS score determined at admission (R^2^ = 0.86, P < 0.001). Patients with complete tSCI at admission had an SPI that was significantly smaller compared to the patient with incomplete tSCI (0.35 ± 0.10 vs. 1.33 ± 0.09; p < 0.001; Fig. 5B). A total of 12 (60.0%) patients showed improvement of their AIS score following their initial assessment (**Table 4**). These patients had a significantly higher SPI compared to patients who did not exhibit AIS score improvement (1.33 ± 0.09 vs. 0.35 ± 0.10, P < 0.001). SPI exhibited a strong positive correlation with the AIS score at last follow-up (R2 = 0.79, p < .001; Fig. 5C). ROC analysis revealed that a SPI cutoff of 1.0 predicted completeness of the spinal cord injury with an area under the curve of 0.99 (P < 0.001; Fig. 5D). Finally, in contrast to our rodent tSCI model, SPI and PAD exhibited only a moderate albeit statistically significant correlation in our patient cohort (R^2^ = −0.59, p < .001; Fig. 5E).

**Fig 5.**
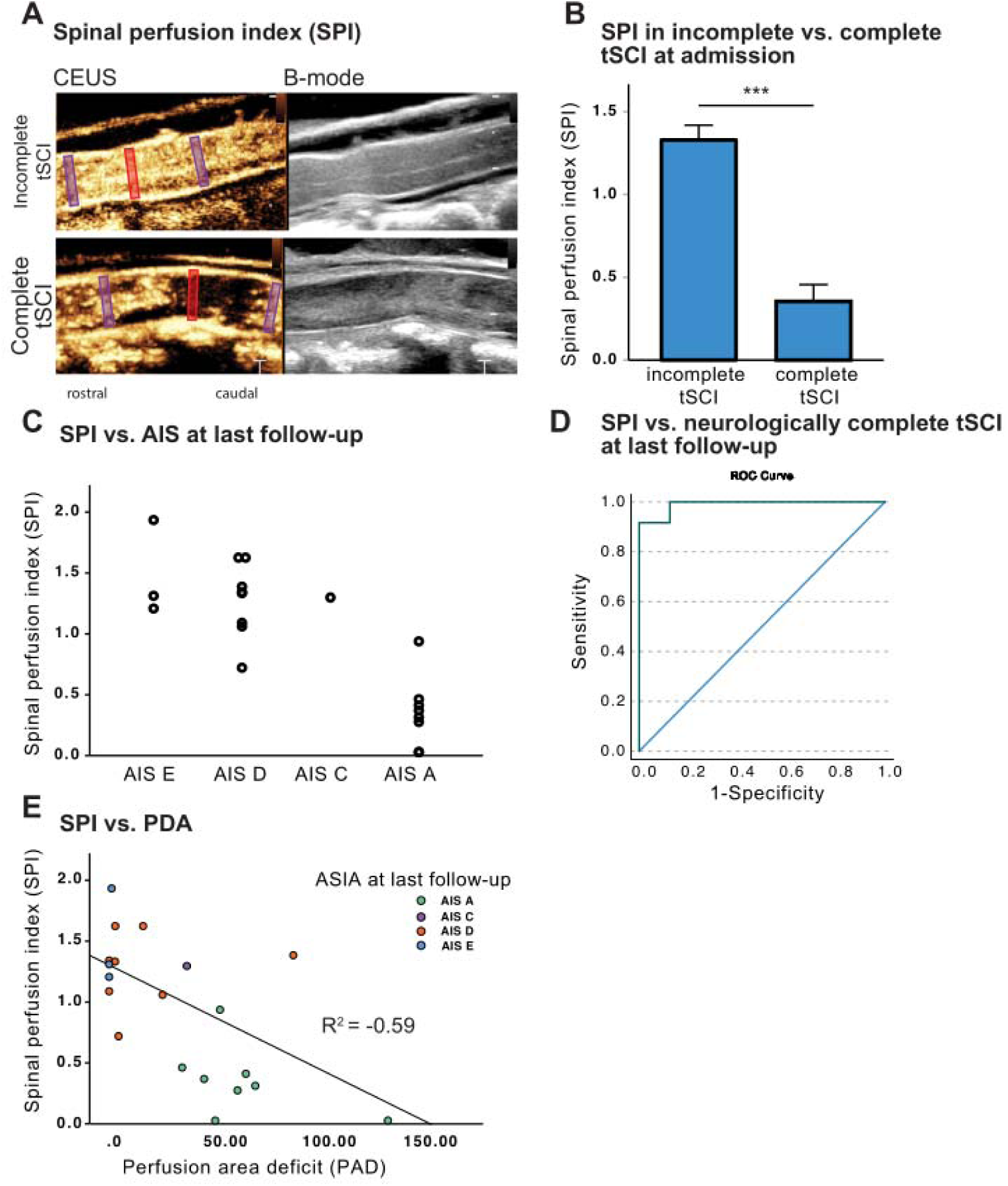
Assessment of spinal perfusion index (SPI) in human tSCI. (A) Representative intraoperative ultrasound displaying tissue perfusion (left panel) and B-mode (right panel) of a motor incomplete (upper row) and motor complete (bottom row) human tSCI. The area under curve within the ROI at the injury center (red) is normalized to the average area under the curve in the rostral (green) and caudal (blue) ROI. Note the striking difference between hyperemia detected in a patient suffering from a motor incomplete tSCI compared to almost complete loss of perfusion in a patient with complete tSCI. (B) Comparison of the SPI between admission exam motor incomplete (ASIA C-E) and motor complete (ASIA A-B) patients. A significantly lower SPI is detected in motor complete tSCI patients. (C) Scatterplot of tSCI patients displaying SPI in relation to ASIA grade at last follow-up. Patients with worse neurological outcomes have a more pronounced perfusion area deficit. (D) Receiver-operating-characteristics of SPI vs. completeness of tSCI at last follow-up. A SPI cutoff of 1.0 resulted in an area under the curve of 0.99 (P < 0.001). (E) SPI and PAD show a moderate negative correlation in our patient cohort.

**Table 4.** Rat Injury Severities.

|  | **Intended Force**<br>**(kDyne)** | **Actual Force**<br>**(kDyne)** | **Number of**<br>**Animals (n)** |
| --- | --- | --- | --- |
| **Mild** | 100 kDyne | 104.13 ± 3.75 | 15 |
| **Moderate** | 150 kDyne | 155.57 ± 4.61 | 7 |
| **Severe** | 200 kDyne | 203.80 ± 2.66 | 10 |

## Discussion

In the current study, we identified CEUS-derived biomarkers to quantify spinal cord damage (PAD) and spared tissue (SPI) in acute tSCI. In rodent models of tSCI, these biomarkers show a strong correlation and have a high predictive value for long-term functional outcomes. The acquisiont of CEUS-derived biomarkers in human tSCI is feasible and revealed distinct patterns of vascular disruption. Notably, the SPI can be readily calculated intraoperatively using the ultrasound console, and its prognostic capacity are promising.

Our proposed biomarker evaluates vascular disruption, a critical intial phathophysiological event following tSCI ^35^. Disruption of spinal cord blood flow with subsequent ischemia has been documented in a variety of tSCI models using techniques such microangiography ^37, 38^, autoradiography ^39, 40^, hydrogen electrode ^41^, and microsphere deposition assay ^38^. In the current report we corroborate the disruption of spinal cord perfusion in our rodent tSCI model using two gold-standard methods: fluorescent microspheres deposition assay and vessel patency using histological methods. Moreover, the disruption of blood flow is directly proportional to the tSCI severity ^42^. Fehlings and colleagues subjected adult rats to mild, moderate, or severe cervical compression injuries. Spinal cord blood flow at the injury site measured with hydrogen clearance technique decreased in a linear fashion with increasing tSCI severity. Our laboratory recently corroborated injury severity-dependent decrease of spinal cord perfusion in rodents using CEUS ^34^. A severe contusion tSCI in the thoracic spinal cord resulted in a significantly larger PAD compared to a moderate tSCI. Importantly, the PAD correlated to the hindlimb locomotor score at 8 weeks post injury. In the current study we demonstrate that the size of the PAD is directly linked to the injury severity in rodent and human tSCI.

However, compared to rodent tSCI, the size of the PAD showed much greater variablity in human tSCI. It was absent in some patients with motor-incomplete injuries while several patients with considerable PAD experienced significant neurological recovery. This difference in the rodent and clinical PAD data is not surprising, given that rodent tSCIs are highly standardized as opposed to the extensive heterogeneity of human tSCI ^43^. Moreover, human tSCI vicitims are of differenent ages, with varying co-morbidities, and pre-existing conditions of the spine such as myelopathy, stenosis, or previous surgeries. Low-energy trauma such as ground level falls can injure the spinal cord especially when it is confined in a stenotic spinal canal (i.e., central cord syndrome) ^44^. Given the aging of our society the incidence of central cord syndrome is rising rapidly ^45^. Additionally, high-energy trauma, such as high-speed car accidents, may result in disruption of the spinal column with violent contusion, stretch, or disruption of the spinal cord. In summary, human tSCI are extremely heterogenous ^46^, which appears to be reflected by different sizes and shapes of the PAD. The heterogeneity of post-traumatic spinal cord tissue perfusion has been previously reported in 22 patients with acute tSCI who underwent intraoperative laser speckle imaging ^47^. The authors identified three distinct pathological patterns of spinal cord blood flow: necrosis-penumbra pattern with low blood flow (necrosis) area at the injury center surrounded by intermediate blood flow (observed in thoracic injuries), the hyperperfusion pattern where blood flow was high throughout the injury site (observed only in cervical injuries), and the patchy-perfusion pattern where low, intermediate, and high blood flow were seen (found in cervical and thoracic injuries). We confirmed the presences of both areas of significant hypoperfusion (a likely necrosis-penumbra pattern as described in Gallagher et al.,) and hyperpefusion pattern using CEUS in our patient cohort. However, the patchy-perfusion pattern pattern was not apparent in our patient cohort. This may be due to our limited patient chort or the fact that this particular perfusion pattern seen in Gallagher et al., is limited to the superficial spinal cord tissue captured by laser speckle imaging while CEUS visualizes the entire diameter of the spinal cord. The impact of various posttraumatic perfusion patterns on injury evolution and recovery is an intriguing topic for future studies.

Our proposed CEUS-derived PAD constitutes a surrogate for damaged spinal cord parenchyma. Interestingly, almost all prognostic biomarkers for tSCI are directly or indirectly related to the amount of damaged spinal parenchyma. Several inflammatory and structural proteins that correlate with the injury severity have been identified in cerebrospinal fluid and serum. The foundational mechanism behind this is the discharge of cellular proteins either into the subarachnoid fluid or the bloodstream from structurally damaged cells. The magnitude of the structural spinal cord damage correlates with injury severity and potentially the clinical outcome ^48^. Among others, inflammatory, autoimmune, neuronal as well as astroglial markers have shown promising results in human trials. Elevated post-traumatic CSF concentrations of cytokines like IL-6 and IL-8, as well as GFAP have been shown to correlate with AIS grades. Notably, protein levels of IL-6, IL-8, MCP-1, tau, S100b, and GFAP are linked with neurological recovery, with reduced levels observed in patients showing improvement ^49^. In addition to that, pNF-H and NSE serve a diagnostic purpose as the serum or CSF concentrations reflect injury severity with elevated levels 24 and 48 hours after the injury ^50, 51^. Furthermore, prolonged pNF-H elevations at 48 hours after the injury have been associated with a higher mortality. However, utilization of serum and CSF biomarkers in the context of spinal cord injuries faces significant limitations. Firstly, their practical application can be hampered by the the need for sample acquisition, particularly in the case of CSF, which necessitates additional labor and time intensive lumbar puncture procedures. Furthermore, these biomarkers require time to become elevated after a spinal cord injury, limiting their effectiveness in acutely sampled CSF, and potentially delaying diagnosis and intervention. Additionally, the variability in sample processing times and the need for standardized laboratory protocols can introduce further complexities in obtaining consistent and reliable results.

MRI-based biomarkers constitute another type of prognostic imaging biomarker for tSCI. An array of sequences has successfully predicted probability of short-term improvements following tSCI. In T2-weighted MRI scans the sagittal intramedullary lesion length, intramedullary hemorrhage, as well as spinal cord signal abnormalities show a strong correlation with the extent of neurologic impairment and functional conversion ^28, 29, 52^. Additionally, MRI abnormalities rostral and caudal to the lesion as well as in the cerebral cortex have been reported to correspond with functional outcomes, suggesting an anterograde and retrograde post-traumatic degeneration ^53–56^. MRI has also been utilized to look at the properties of spared spinal cord parenchyma. The utilization of diffusion tensor imaging (DTI) MRI presents a promising method to assess white matter pathology in tSCI ^57, 58^. DTI analyses the speed and direction of water diffusion to assess fiber tracks microstructure and anatomy and is feasible in human tSCI ^59^. However, the use of MRI biomarkers in spinal cord injuries faces notable limitations. First, correlation of MRI-based fiber tractography with neurological impairments has been only demonstraed in chronic tSCI ^60^. DTI obtained acutely after tSCI lack prognostic capacity since potentially preseverved white matter tracts are distracted and compressed by the disrupted spinal column ^61^. Second, the spatial resolution of MRI can be inadequate for capturing fine structural changes which indicate tissue damage. Third, not all acute tSCI patients undergo MRI for various reasons: the unavailabilty of MRI during off hours, the limited value of MRI on treatment decisions, or the patients being medically too unstable. In our level 1 trauma center only approximately 50% of tSCI patients undergo pre-operative MRI. These challenges underscore the urgent need to develop alternative diagnostic imaging methods.

We advocate for the incorporation of contrast-enhanced ultrasound (CEUS) as a cutting-edge imaging modality for evaluating acute tSCI. This technology offers a host of compelling advantages compared to other biomarkers. Foremost, its remarkable spatial resolution capabilities (∼100 micrometer) enable precise characterization of injury extent and identification of areas with spared spinal cord parenchyma. Real-time ultrasound imaging provides surgeons with immediate access to dynamic physiological information, motivating additional surgical maneuvers in approximately one third of our patients. Of particular significance is the technology’s ability to assess perfused and viable tissue at the injury site intraoperatively. Spared perfused spinal cord tissue is the target of neuroprotective interventions and serves as a predictor of functional outcomes.

The following limitations of our study should be noted: The proposed CEUS biomarker can only be obtained during posterior decompression surgery since the optical window is not large enough to allow for imaging via anterior approaches. However, this limitation has become less prominent, since posterior spinal cord decompression is recommended in the setting of acute tSCI as it allows for more thorough spinal cord decompression and has been associated with improved neurological outcomes ^62^. Another challenge lies in the operator-dependent nature of ultrasound imaing, especially since tSCI patients often present during non-standard hours with a lack of imaging staff. Therefore, training spine surgeons in the acquisition of intraoperative CEUS becomes crucial to enhance the accessibility of this technique with growing interest^63^. Lastly, to further elucidate the prognostic capabilities of PAD and SPI in human tSCI, larger multicenter studies are imperative, aiming to provide a more comprehensive understanding of these biomarkers’ potential.

In conclusion, CEUS-derived acute biomarkers allow for the estimation of injury severity by quantifying the spared perfused spinal cord parenchyma at the injury center of both rodent and human tSCI. The proposed quantitative biomarker for spared tissue (SPI), displays a remarkable prognostic capacity in both animal and human tSCI. Future studies are needed to further refine the standard operating procedures for acquisition and analysis of CEUS imaging for optimal prognostic capacity.

## Materials and Methods

### Animal Use & Spinal Cord Injury Model

All animal procedures were performed in accordance with the guidelines from Office of Animal Welfare from the National Institutes of Health and approved by the University of Washington Institutional Animal Care and Use Committee (IACUC Protocol # 4362-01). Animals were randomly assigned to different treatment groups and researchers were blinded to their status during the behavioral studies and analyses. Female Long Evans rats (n= 32, 8-12 weeks old; weight 250 – 300 grams, Envigo Labs) were anesthetized using isoflurane (5% to induce and 2.5% to maintain), shaved and the area overlying the T6 - T11 vertebrae was cleaned and sterilized. Using sterile techniques, an approximately 2.5cm longitudinal incision centered over T7 – T10 was made before subperiostal dissection of paraspinal muscles. A laminectomy was performed to expose the spinal cord at T8. To produce a contusion type lesion at T8, an Infinite Horizon Device Impactor (IH-0400, Precision Systems and Instrumentation, LLC, Lexington KY) equipped with a 2.5mm diameter probe tip. Three different injury severities were produced for this study: mild (100 kDyne; n = 15), moderate (150 kDyne; n = 7) and severe (200 kDyne; n = 10) injuries. Tissue displacement and peak force data were collected, and actual force recorded was used as indicators of injury consistency and severity (see **Table 4**). Post-surgical animals were monitored twice daily for dehydration and general health and manual bladder expression was performed up to 14 days post injury (dpi). Spontaneous voiding reflexes returned between 7 – 10 dpi. To manage pain associated with the surgical procedure, the animals were injected with buprenorphine (0.03 mg/kg) subcutaneously every 8-12 hours for 48 hours after injury. Antibiotics were administered daily for 5 dpi as infection prevention (gentamicin, 5mg/kg Pfizer, Inc.).

#### Ultrasound Imaging and Analysis in Rats

The Vantage ultrasound research platform (Verasonics, Kirkland, WA, USA) was used to program multi-angle plane-wave nonlinear Doppler sequences, using a 15MHz linear array transducer (Vermon, France). The ultrasound probe is placed approximately 5mm above the dorsal spinal cord surface over the midline sagittal plane and warmed sterile ultrasound gel acoustically coupled the transducer to the spinal cord. For each contrast injection, a bolus of 0.lmL of Definity® (Lantheus, New Jerssey, US) was injected intravenously followed by a 0.2mL of saline flush. Multi-angle plane wave transmits are acquired at 500-2000 Hz pulse repetition frequency were collected. After beam forming and IQ demodulation, ensembles were filtered using a singular value decomposition (SVD)-based approach to segment flow signals based on relative velocity. This results in “vascular flow” and “perfusion” image ensembles, to be analyzed independently ^64, 65^. Image acquisition was initiated prior to contrast inflow and will proceed for 30 seconds after the peak. Short nonlinear ensemble acquisitions (40 frames, amplitude modulation pulsing scheme) was conducted at an inter-ensemble frame rate of 16Hz. This pulsing scheme eliminates tissue signal, and the use of short, repeated ensembles facilitates the isolation of microcirculatory flow signal from corrupting larger vascular signal using an adaptive spatiotemporal filtering approach. Quantitative parameters were extracted from the resulting time-intensity curve (TIC) for each spatial voxel to generate a parametric output image.

Two quantitative measurements, perfusion area deficit (PAD) and spinal perfusion index (SPI), were obtained from CEUS data acquired at midline longitudinally. Custom Matlab scripts were written to quantify both metrics. A region of interest (ROI) was manually drawn over the hypoperfused area on CEUS Doppler images to calculate PAD. Three rectangular ROIs were drawn on area under the curve (AUC) maps derived from CEUS bolus acquisitions to calculate SPI (one rostral, one at epicenter, one caudal; 1 mm wide x 3 mm tall, spaced 3 mm apart). The average signal at the epicenter was divided by the average signal in the rostral and caudal ROIs to obtain SPI. Only signal within the cord was included for analysis (i.e., signal from vertebrae and meninges was excluded).

#### Behavioral Assessment

Animals were acclimated to being handled and to the various pieces of behavioral equipment that they would be tested in for at least two weeks before surgery. Baseline behavioral recording for all tests was done at one week prior to surgery.

### Open-fiend Locomotor Test (Basso, Beattie and Breshnahan – BBB test)

The BBB locomot or rating scale is a sensitive, observer-scored test of hindlimb locomotor function that is commonly used in rodent model after SCI ^66^. Animals are scored according to a standardized score sheet from 0 (no hindlimb movement) to 21 (complete locomotor recovery). Animals are placed into a ∼100 cm diameter arena with 19 cm walls (a children’s play pool) that has a lightly textured, uniform floor. For four minutes, each animal is permitted to explore the arena while two observers record locomotor actions according to the BBB Locomotor Rating Scale ^66^. The observers compare scoring, which was done live, and, if necessary, come to a consensus on the final left and right hindlimb scores. The BBB test was repeated every week for eight weeks. The test was video recorded for reviewing and confirmation of scores if needed.

### Ladder Walk

The ladder walk test requires no pre-training. This test evaluates sensory-induced motor function of the rodent’s limb ^67^. The apparatus consists of two clear plexiglass walls ∼90 cm long and ∼15 cm tall that are ∼10 cm wide, creating a corridor. A series of 1 mm diameter stainless steel rungs are intermittently spaced (gaps range between 1 - 3 cm) to create a ladder-type floor. The entire apparatus is elevated slightly (∼ 30 cm) to create a space below for the animal’s leg to fall. The home cage was placed at one end of the apparatus. For each trial, the animal is placed at the end opposite to the home cage and walks the length of the apparatus to return to the home cage. Between each successful run, the placement of the rungs was changed to prevent the animals from memorizing a specific rung pattern. The test consists of ten trials and was recorded on video. The videos were later reviewed at half speed; the total number of attempted steps and the number of slips or misses (unsuccessful grasps of the rung) were counted for each hind limb. The percentage of errors was recorded. The animals were tested prior to injury to establish a baseline error percentage; then again at 4- and 8-weeks post injury (wpi).

### Histological Analysis for Blood Vessel Patency

To assess intrinsic spinal cord blood vessel patency, we used fluorescein isothiocyanate (FITC)-conjugated *Lycopersicon esculentum agglutinin* (tomato) lectin (LEA, Sigma-Aldrich). Prior to tSCI, 0.2 mL (1 mg/mL) are injected via a tail-vein. Following tSCI animals receive a second injection of Texas-red labeled lectin. Following 30 minutes, animals are euthanized. A 10 mm long segment centered over T8 is analyzed for density of patent microvasculature. For quantification a grid of 500 µm squares is placed over longitudinal photomicrographs using ImageJ. Within each square, distinct microvascular fragments were counted using the multi-point counter tool in ImageJ.

### Fluorescent Microsphere Deposition Assay

Following thoracic tSCI rats were positioned in the supine position and an approximately 1 mm incision was made on the right side of the neck. Blunt dissection was carried out to identify and isolate the carotid artery. Heparinized PE50 tubing was introduced into the artery and advanced to the heart. An in-line pressure monitor was used to confirm the tubing location in the right atrium. 4-0 silk sutures were used to secure the tubing in place. Red or yellow, fluorescent microspheres were randomly chosen as either the pre- or post-injury color (Invitrogen™; F8842, F21011). The microspheres were vortexed immediately before injection into the right atrium (0.5mL). The animal was then prepared for and given the spinal cord injury, as described above, before the post-injury fluorescent microsphere injection of the alternate color (0.5mL). Animals were euthanized approximately 30 min after the microsphere injection. Spinal cord tissue (unfixed) was collected. A 10mm long spinal cord segment centered at the injury was sectioned 16µm thick on an Imaging Cryomicrotome (Barlow Scientific, Inc, Olympia, WA) ^68^. Fluorescent microsphere density was calculated using in-house scripts written for ImageJ and MATLAB (R2017, Mathworks Inc; Natick, MA).

### Histological Analysis for Spared Tissue

Animals were sacrificed at 8 wpi with an overdose of pentobarbital sodium (intraperitoneal, 0.39g/mL Schering-Plough Animal Health, Madison, NJ). Animals were fixed via trans-cardiac perfusion with 200mL of ice-cold PBS (pH 7.4) followed by 200mL of 4% paraformaldehyde (PFA). Spinal cords were harvested and post-fixed in 4% PFA at 4°C for 16-18 hours. Spinal cords were mounted and sectioned cross-sectionally using a cryostat (CM1850, Leica Biosystems) at 20μm thickness. Sections were collected, and thaw mounted onto gelatin-coated slides and stored at −80°C until use. To perform staining, sections were thawed and fixed with 4% PFA (Macron Chemicals, Center Valley, PA) for 15 minutes to ensure tissue adherence. Sections then underwent a series of alcohol incubations for dehydration beginning with 4 min in deionized water, followed by 5 min periods in 70%, 95%, and 100% ethanol following with a xylene incubation for 10 min and then back through the ethanol series. Sections were then incubated for 10 minutes in an aqueous myelin dye (Eriochrome Cyanine R; Fluka, St. Louis, MO) and differentiated for 1 min in 1% ammonium hydroxide (Thermo Fisher Scientific, Fair Lawn, NJ) after washing in water. Sections were then briefly rinsed in water then incubated in Cresyl violet stain (Sigma-Aldrich, St. Louis, MO) for 3 minutes. Following this step, sections were rinsed with deionized -water 10 times and then quickly dehydrated in a stepwise manner for 4 min each in 70% and 95% alcohol. The Cresyl violet was then briefly differentiated in a 250mL of 95% alcohol solution containing five drops of glacial acetic acid (Thermo Fisher Scientific). Once differentiation was sufficient, sections continued through a stepwise dehydration process consisting of 5 min each in 95% and 100% ethanol, followed by final 10-minute incubation in xylene. Slides were then cover slipped using Permount. CV/myelin-stained tissue sections (sampled at 250μm interval for 7mm in length) were imaged using a Zeiss Primo Star microscope with a color camera (Axiocam ERc 5s).

### Intraoperative Ultrasound in Patients with Acute tSCI

Patients who presented to the Harborview Medical Center emergency room within 72 hours after acute tSCI were screened for inclusion in our study. Patients met criteria for inclusion if they were at least 18 years of age, had a neurological deficit on examination (ASIA A-D), and were medically stable to undergo routine posterior spinal decompression and stabilization. Patients were excluded if they had co-existing TBI or penetrating injuries. A study protocol outlining the recruitment of patients, data collection, data storage and use was reviewed and approved by the University of Washington IRB. Data was collected pseudonymously after written and informed consent in accordance with national law and the 1975 declaration of Helsinki. Upon completion of a routine posterior decompression and stabilization CEUS was performed in the operating room at Harborview Medical Center. A Philips EPIQ premium ultrasound system in combination with an intraoperative ultrasound probe (L12-3) was used. First, we obtained B-mode midline sagittal images of the entire rostrocaudal extent of the exposed spinal cord. Prior to CEUS, the contrast agent (Definity®, Lantheus Medical Imaging) was activated in a full 45-second activation cycle using a VIALMIX ® activator and diluted in saline (1.3ml Definity/8.7ml saline) ^69^. Using a peripheral intravenous cannula, a weight-adjusted bolus injection was delivered (10µl Definity/kg) while the ultrasound transducer was positioned in a midline sagittal plane. Data acquisition was initiated at the time of contrast injection and continued for up to 3 minutes following injection to capture the full dynamic enhancement curve.

### Analysis of Intraoperative Ultrasound Images

CEUS images were analyzed utilizing the Phillips Q Lab software. First, we utilized the area outline tool to calculate the size of the perfusion area deficit on CEUS images (PAD). To assess the amount of contrast inflow into the spared tissue at the injury center, we placed a rectangular region of interest (ROI, 3mm wide and spanning the diameter of the spinal cord, Fig 5) onto the injury center. Contrast-inflow into intact cord tissue was assessed by pacing duplicate ROIs rostral and a caudal to the damaged spinal cord areas as determined with B-mode and contrast inflow images. The contrast intensity curve was calculated by the software. The spine perfusion index (SPI) was calculated by normalizing the AUC at the injury center with the average AUC obtained from intact spinal cord rostral and caudal to the injury. The SPI was calculated using the following formula: center perfusion/mean peripheral perfusion.

### Statistical Analysis

We analyzed our data using Excel (Microsoft; Redmond, WA), Prism 7 (GraphPad Software; San Diego, CA) and SPSS (28.0, IBM). Continuous variables are shown as means ± standard deviation. To identify significant differences between the groups, we performed the non-parametric Mann-Whitney U Test. Correlative analysis for CEUS biomarker, tissue sparing, and functional outcomes were carried out using the Spearman correlation test.

## Data Availability

All data produced in the present study are available upon reasonable request to the authors.

## List of supplementary Figures

**Fig. S1.**
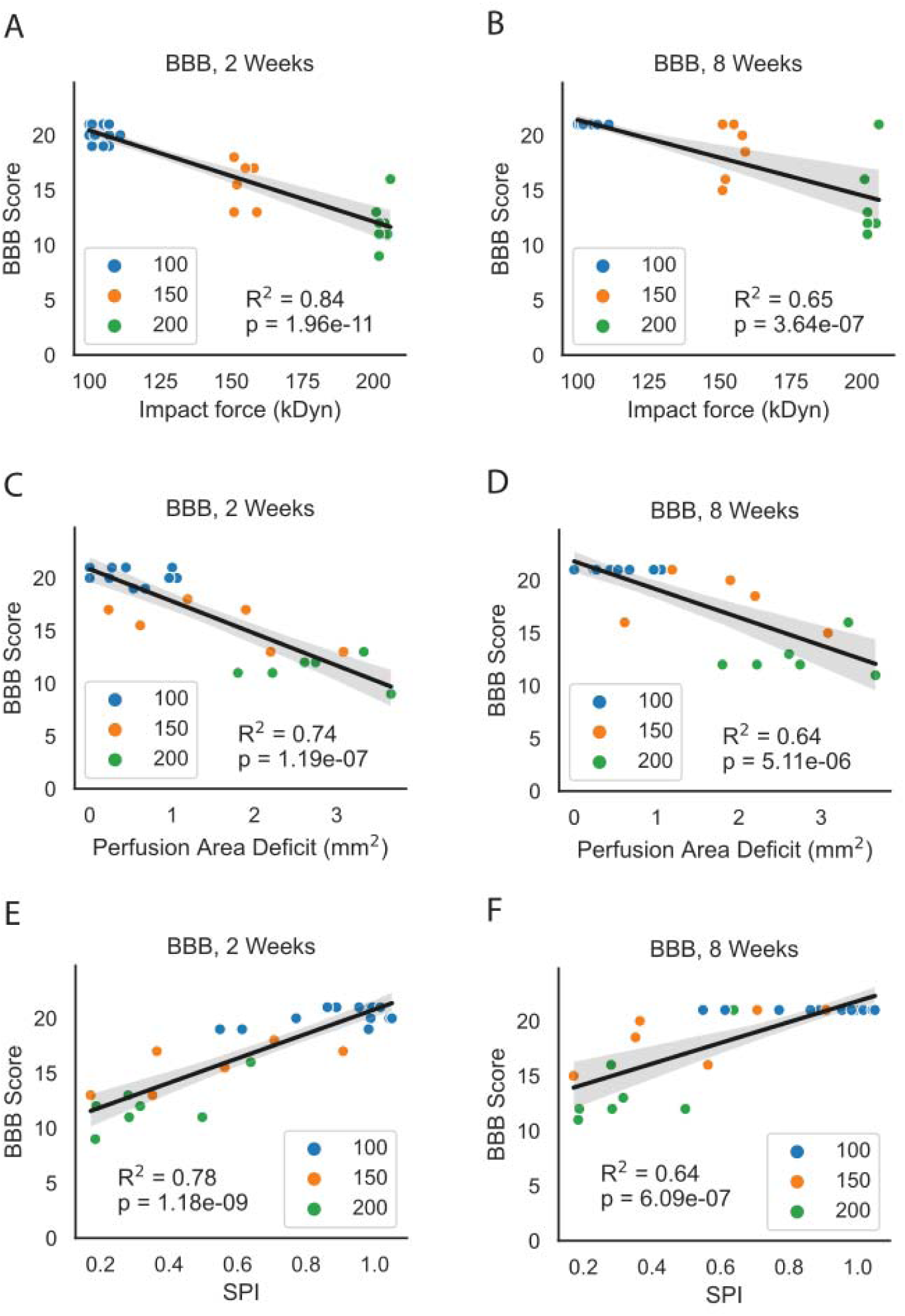
Contrast enhanced ultrasound imaging-based measurements correlate with behavioral outcomes. Perfusion area deficit (PAD) and spinal perfusion index (SPI) were obtained from acute CEUS images of animals with mild, moderate, and severe injuries. Open-field behavioral test (BBB) was performed at 2- and 8-weeks post injury (wpi) R2 =0.84 with p <0.0001; R2 = 0.65 with p<0.0001 respectively). A – B. PAD correlated significantly with BBB scores at 2 and 8wpi with R^2^ =0.84 with p <0.0001 and R^2^ = 0.65 with p<0.0001 respectively. C – D. Spinal perfusion index (SPI) correlated significantly with BBB scores at 2 and 8wpi with R^2^ =0.74 with p <0.0001 and R^2^ = 0.64 with p<0.0001 respectively. E – F. Actual forces obtained from the Infinite Horizon device were correlated with BBB scores at 2- and 8-wpi with R^2^ =0.78 with p <0.0001 and R^2^ = 0.64 with p<0.0001 respectively.

**Fig. S2.**
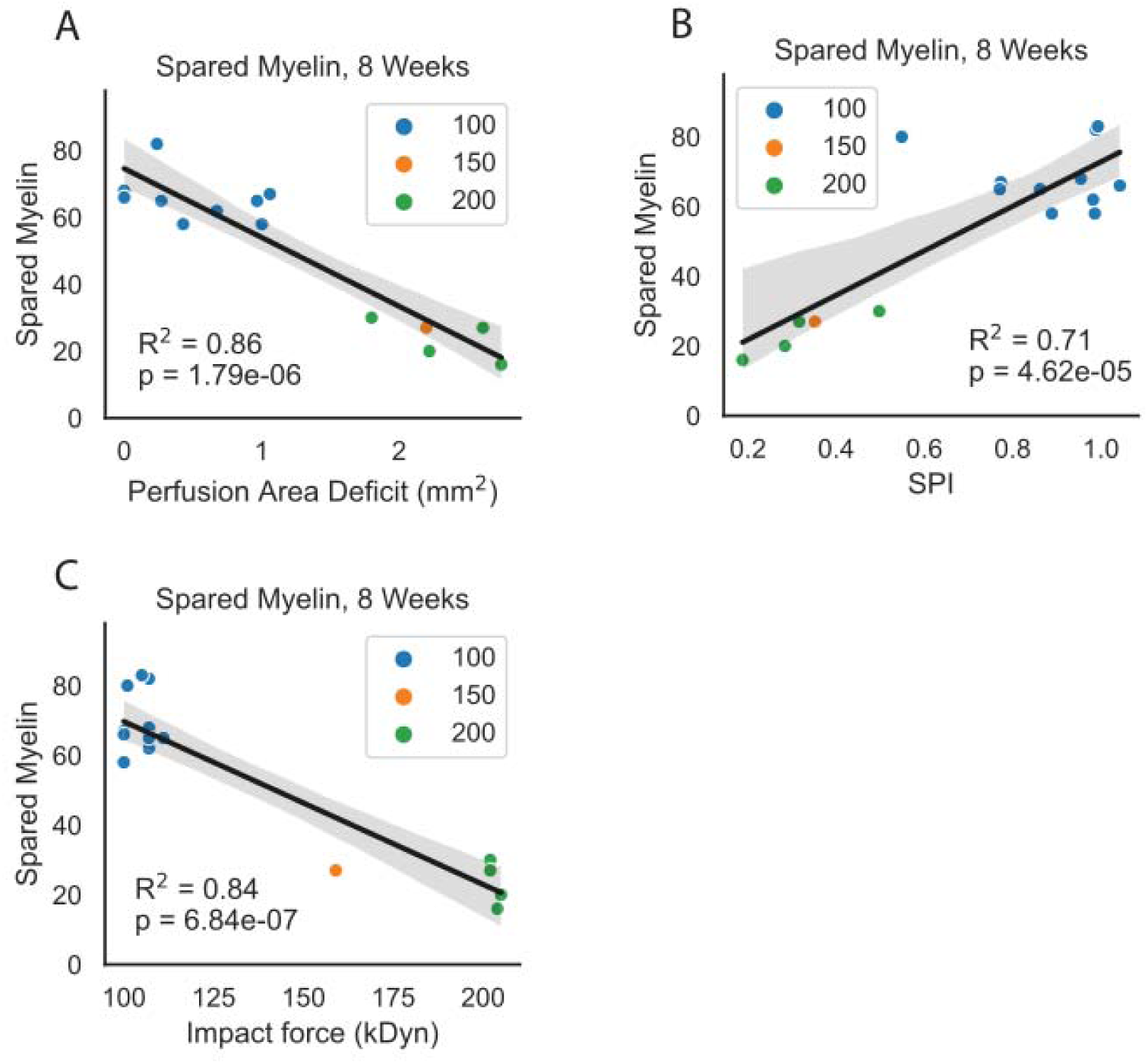
Contrast enhanced ultrasound imaging-based measurements correlate with tissue sparing. Perfusion area deficit (PAD) and spinal perfusion index (SPI) were obtained from acute CEUS images of animals with mild, moderate, and severe injuries. Spinal cord sections stained with Cresyl violet and myelin at every 200 microns were used to determine tissue sparing at 8 weeks post injury. A. PAD correlated significantly with tissue sparing at 8 wpi (R^2^ = 0.86 with p <0.0001). B. Spinal perfusion index (SPI) correlated significantly with tissue sparing at 8 wpi (R^2^ = 0.71 with p <0.0001). C. Actual forces obtained from the Infinite Horizon device correlated significantly with tissue sparing at 8 wpi (R^2^ = 0.84 with p <0.0001).

## Funding

This work is supported by the Department of Neurological Surgery, Royalty Research Fund at the University of Washington, Washington State Spinal Cord Injury Fund, Lantheus (providing microbubbles for the rodent studies), the Craig Neilsen Foundation (#546901 to CPH and ZZK), the Department of Defense (CDMRP TRA-W81XWH-18-1-0753 to CPH and ZZK), and NIH NINDS R01-NS121191 (to ZZK).

## Author contributions

Conceptualization: ZZK, CPH

Methodology: ZZK, CPH, JNH, MK, RWG, MB

Investigation: ZZK, CPH, JL, JNH, SS, LNC, JEH, MK, RWG, MB, CPH

Visualization: ZZK, LNC, JEH, MK, RWG, CPH

Project Administration: ZZK, CPH

Supervision: ZZK, CPH

Writing: ZZK, JL, CPH

Writing – review & editing: ZZK, JL, JNH, SS, LNC, JEH, MK, RWG, MB, CPH

## Competing interest

The authors declare that they have no competing interests.

## Notes

### Competing Interest Statement

The authors have declared no competing interest.

### Clinical Trial

NCT04056988

### Author Declarations

A study protocol outlining the recruitment of patients, data collection, data storage and use was reviewed and approved by the University of Washington IRB. Data was collected pseudonymously after written and informed consent in accordance with national law and the 1975 declaration of Helsinki.

